# Estimation of COVID-19 transmission rates in California and the U.S. with reporting delays

**DOI:** 10.1101/2020.05.14.20101162

**Authors:** Lee Worden, Rae Wannier, Micaela Neus, Jennifer C. Kwan, Alex Y. Ge, Eugene T. Richardson, Travis Porco

## Abstract

We estimated time-varying reproduction numbers of COVID-19 transmission in counties and regions of California and in states of the United States, using the Wallinga-Teunis method of estimations applied to publicly available data. The serial interval distribution assumed incorporates wide uncertainty in delays from symptom onset to case reporting. This assumption contributes smoothing and a small but meaningful increase in numerical estimates of reproduction numbers due to the likely existence of secondary cases not yet reported. Transmission in many areas of the U.S. may not yet be controlled, including some areas in which case counts appear to be stable or slowly declining.

## Introduction

The global COVID-19 pandemic caused by the novel coronavirus SARS-CoV-2 was first confirmed to have arrived in California on January 26, 2020,^1^and community transmission in California was first reported on February 26.^2^Since that time, the disease has caused over 69,000 cases and over 2,800 deaths in California,^3^and over 1.3 million cases and 79,000 deaths in the U.S.^4^The U.S. government declared a public health emergency on January 31, and many states and counties have introduced control measures such as shelter in place orders.

In San Francisco, California and six other nearby counties including Santa Clara County, an official order to shelter in place^5^ took effect on March 17, 2020, directing people to stay at home and avoid all but essential gatherings, travel, and business. Schools and some day care facilities and public places such as parks have additionally been closed. On March 19, a similar stay-at-home order was issued covering the entire state of California.^6^ More recently, San Francisco is introducing a program of contact tracing, and has begun to require the use of cloth face masks beginning April 18, 2020.^7^ At the time of this writing, the shelter-in-place order has been extended through the month of May. In many places, the accumulation of new cases per day appears to have slowed, suggesting that interventions such as social distancing may be helping to slow transmission.

We assessed changes in transmission rates over time by estimating time-varying reproduction numbers. A time-varying reproduction number, commonly written *R_t_*, denotes the average number of cases infected by a given case over the course of that individual’s disease progression, indexed by a time variable *t*, such as the date of the symptom onset of the case. Of particular interest is the question of whether transmission is supercritical, meaning that *R_t_* > 1, in which case the epidemic can increase in size, or is subcritical, meaning that *R_t_* < 1, in which case it will fade out. To eliminate a disease locally, it is not necessary to reduce *R_t_* to zero, only to reduce it below one for a sustained period. We used publically available daily counts of COVID-19 cases by county and state to estimate the time course of the reproduction number in the 50 U.S. states and in counties and regions of California.

## Methods

Daily counts of confirmed cases by state and county have been published by The New York Times.^8^ We used daily reported case counts to estimate effective *R* by day in California counties and in US states, using the Wallinga-Teunis technique (Cauchemez et al. 2006) of real-time estimation.

This technique requires an estimate of the disease’s serial interval distribution, the time in days from the date of reporting of a primary case to that of a secondary case caused by the primary. We estimated the serial interval using estimates of the incubation interval, timing of transmission relative to symptom onset, and timing of case reporting relative to symptom onset, as follows. The incubation interval was modeled as log-normally distributed with mean 5.6 days and standard deviation 4.2 days (logmean 1.5, logSD 0.67) (He et al. 2020). The interval from primary cases’s symptom onset to a transmission event, which may be negative due to presymptomatic transmission, was modeled by 2.5 days plus a gamma distributed number of days with scale parameter 1.5 and shape parameter 2.1. With this assumption, about 44% of transmission is presymptomatic (He et al. 2020). The interval from symptom onset to case reporting, which contributes variance but no difference in mean to the serial interval distribution, was included to reflect variability in reporting, using a log-normal distribution with mean 8.9 days and standard deviation 7 days (logmean ln 7, logSD ln 2), to reflect the possibility that some cases may be detected soon after symptom onset, while others may be detected several weeks later. The resulting serial interval distribution has mean 6.3 days and standard deviation 10.9 days (Figure 1).

**Figure 1:**
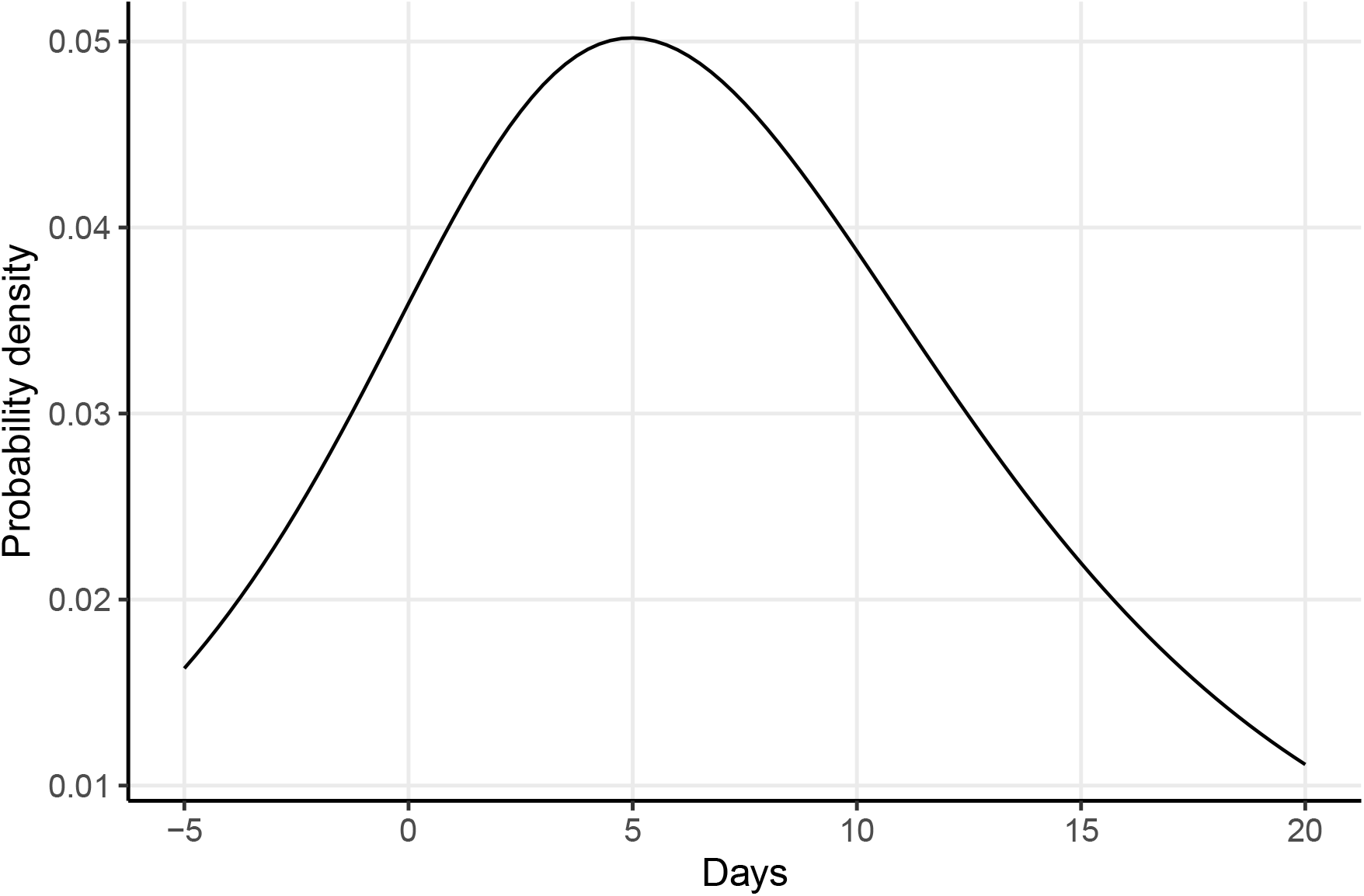
Estimated serial interval distribution.

We applied the Wallinga-Teunis technique to county-level data in California, and to aggregate counts from each of the U.S. states. Counties where cases are relatively sparse were aggregated into regional counts (see Appendix). We then estimated the current reproduction number in each of these localities by combining the daily estimates over the most recent week, using multiple imputation (Rubin and Schenker 1991).

## Results

Daily counts of COVID-19 cases were analyzed by county in the San Francisco Bay Area (Figure 2), by county and region in the rest of California (Figure 3), and by state for the entire U.S. (Figure 7).

**Figure 2:**
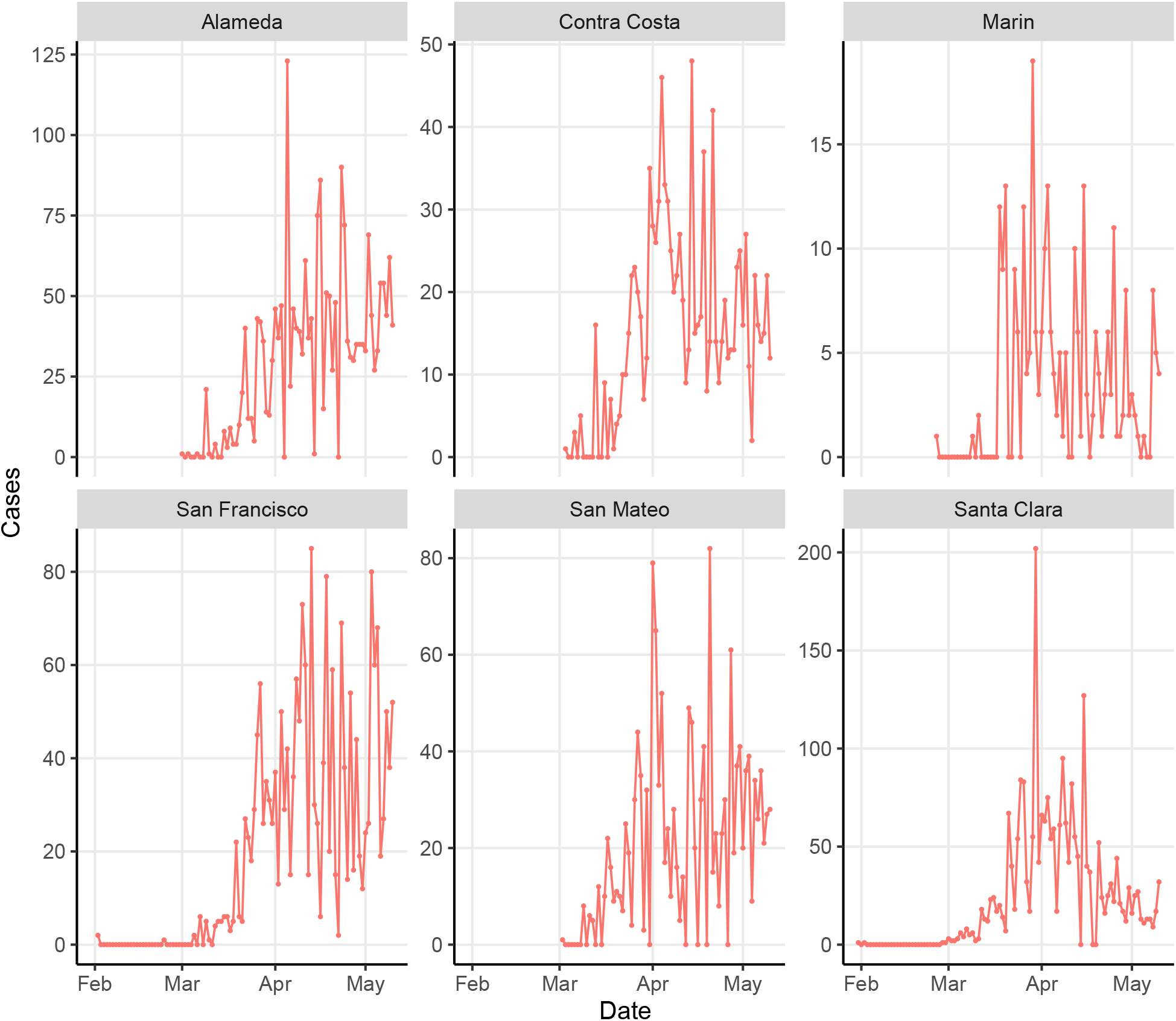
Daily counts of new cases by county in the San Francisco Bay Area

**Figure 3:**
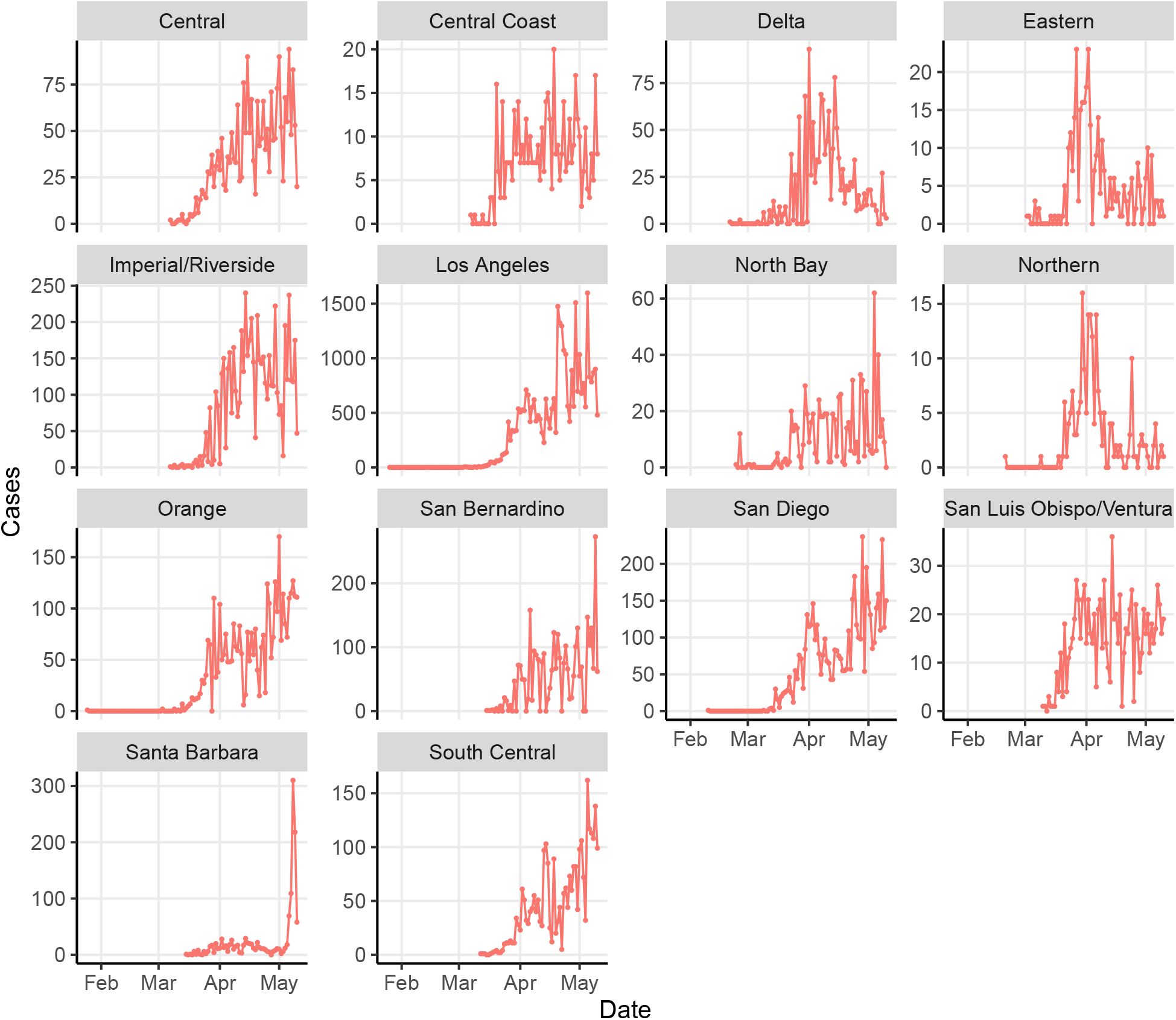
Daily counts of new cases by county or region in California outside of the San Francisco Bay Area

Many counties and regions across California display a characteristic pattern in which estimated reproduction numbers initially rise to a supercritical value then fall to a lower value, roughly coinciding with times before and after the introduction of various control measures in March and April such as California’s stay-at-home order of March 19th (Figures 4, 5). A large increase in cases in early May was seen in Santa Barbara County, likely reflecting cases identified in Lompoc Prison.^9^

**Figure 4:**
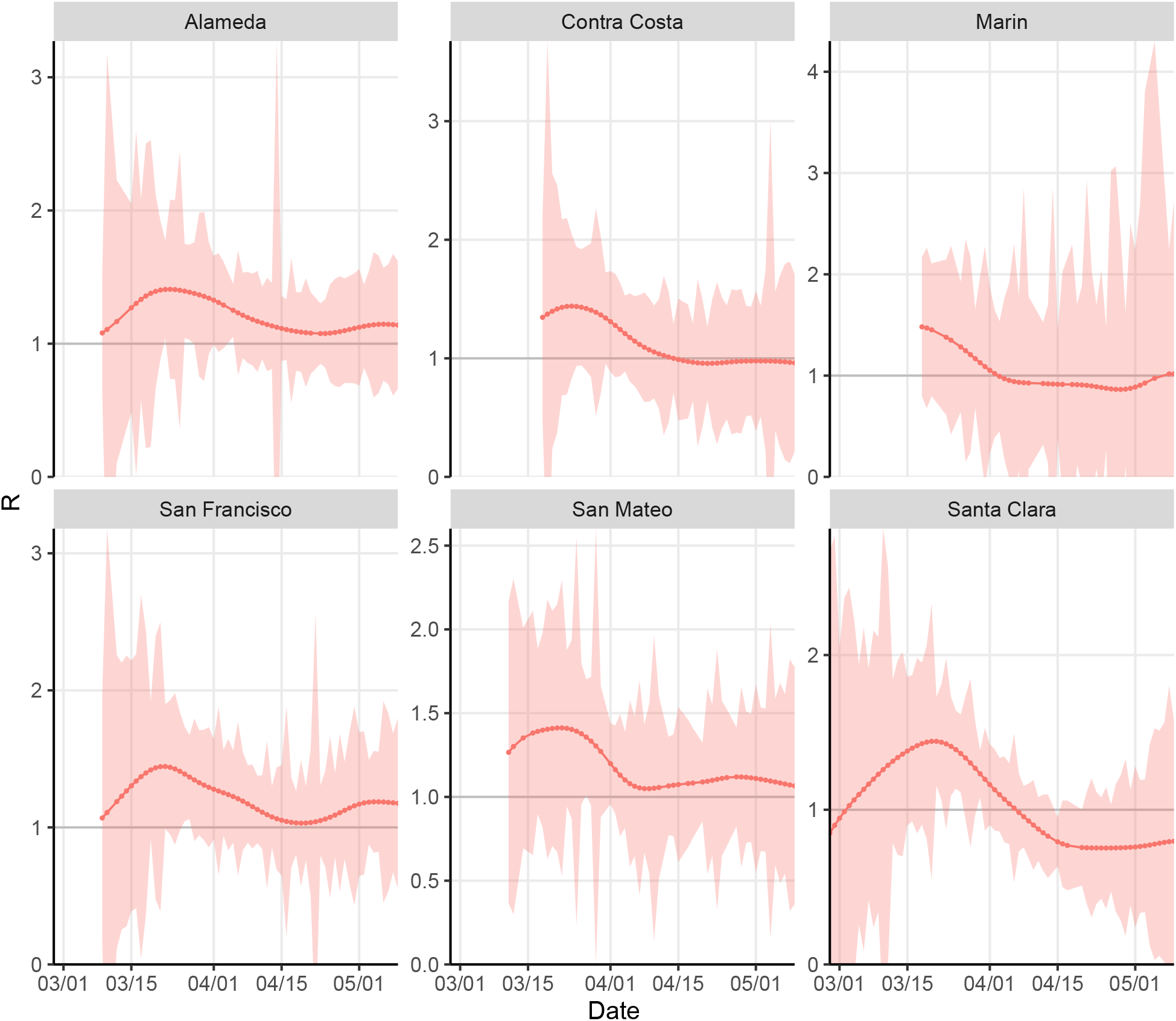
Estimated effective *R_t_* by county in the San Francisco Bay Area

**Figure 5:**
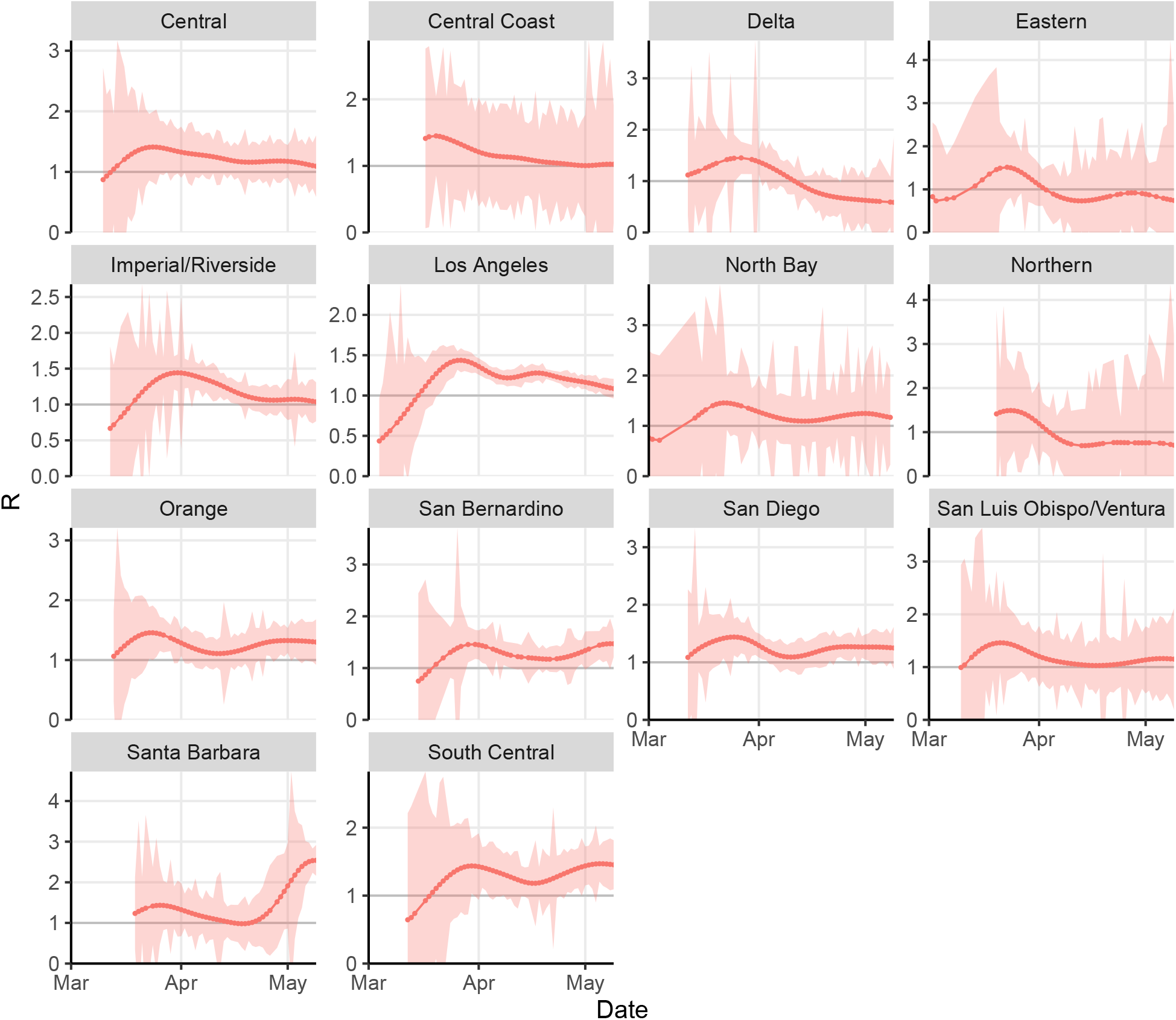
Estimated effective *R_t_* by county or region in California outside of the San Francisco Bay Area

A similar pattern is observed in many states (Figure 8), while in others *R_t_* remains high, reflecting that daily case counts are still rising. In particular, our central estimate of *R_t_* in New York became subcritical (*R_t_* < 1) on approximately April 12, 2020, while this occurred in New Jersey on approximately April 21, 2020. As of May 9, 2020, the estimated *R_t_* for Minnesota was still above 1.5, while for Nebraska, the value was about 1.34. The estimated *R_t_* in South Dakota as of May 9, 2020 was also approximately 1.5, this estimate being influenced by testing in Minnehaha County which brought over 200 cases to light.

Estimates of most recent *R_t_* (Figures 6, 9) confirmed that values range from subcritical to supercritical, with many confidence intervals spanning the critical threshold of one.

**Figure 6:**
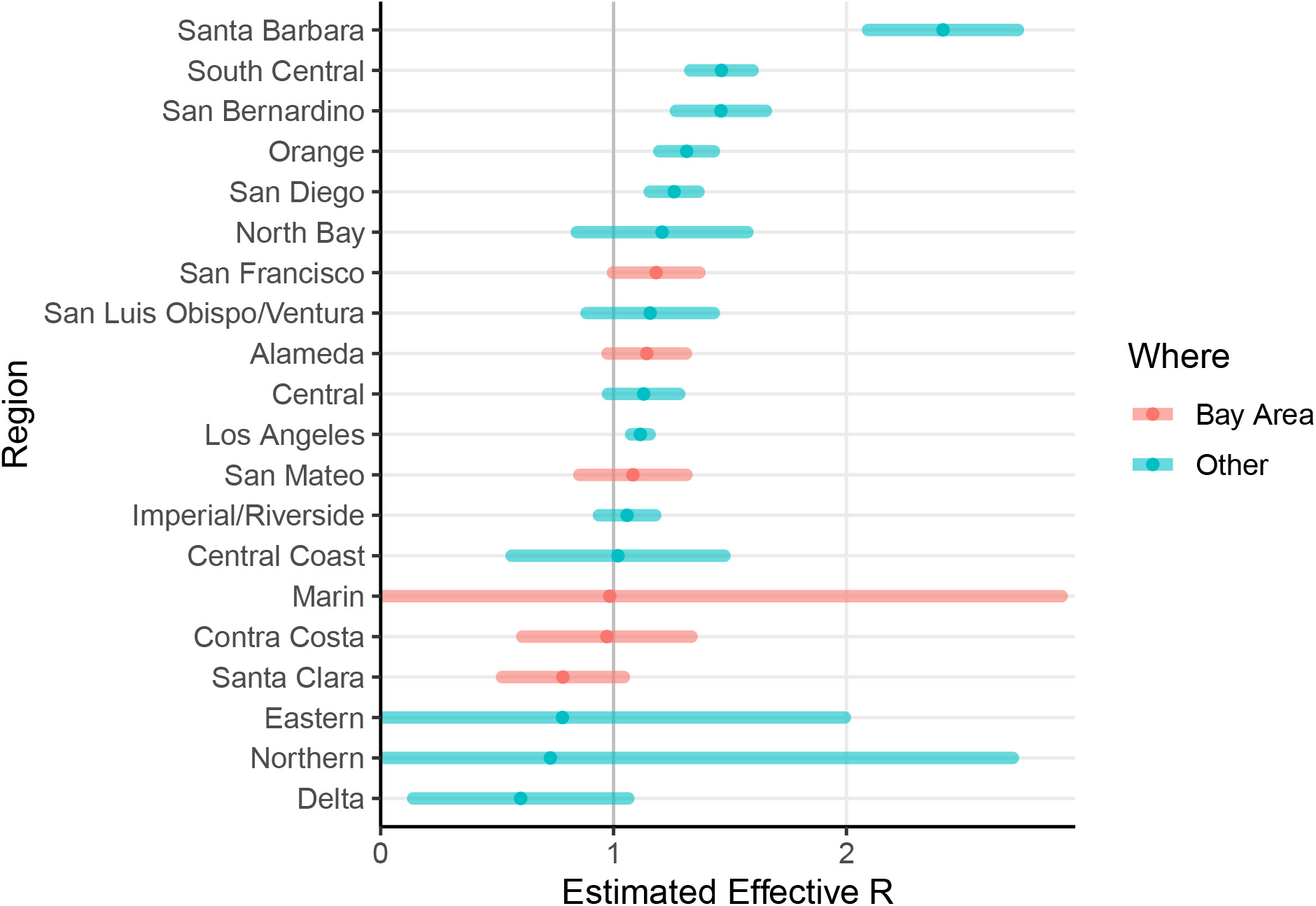
Most recent estimates of effective *R_t_* by county or region in California, in descending order.

**Figure 7:**
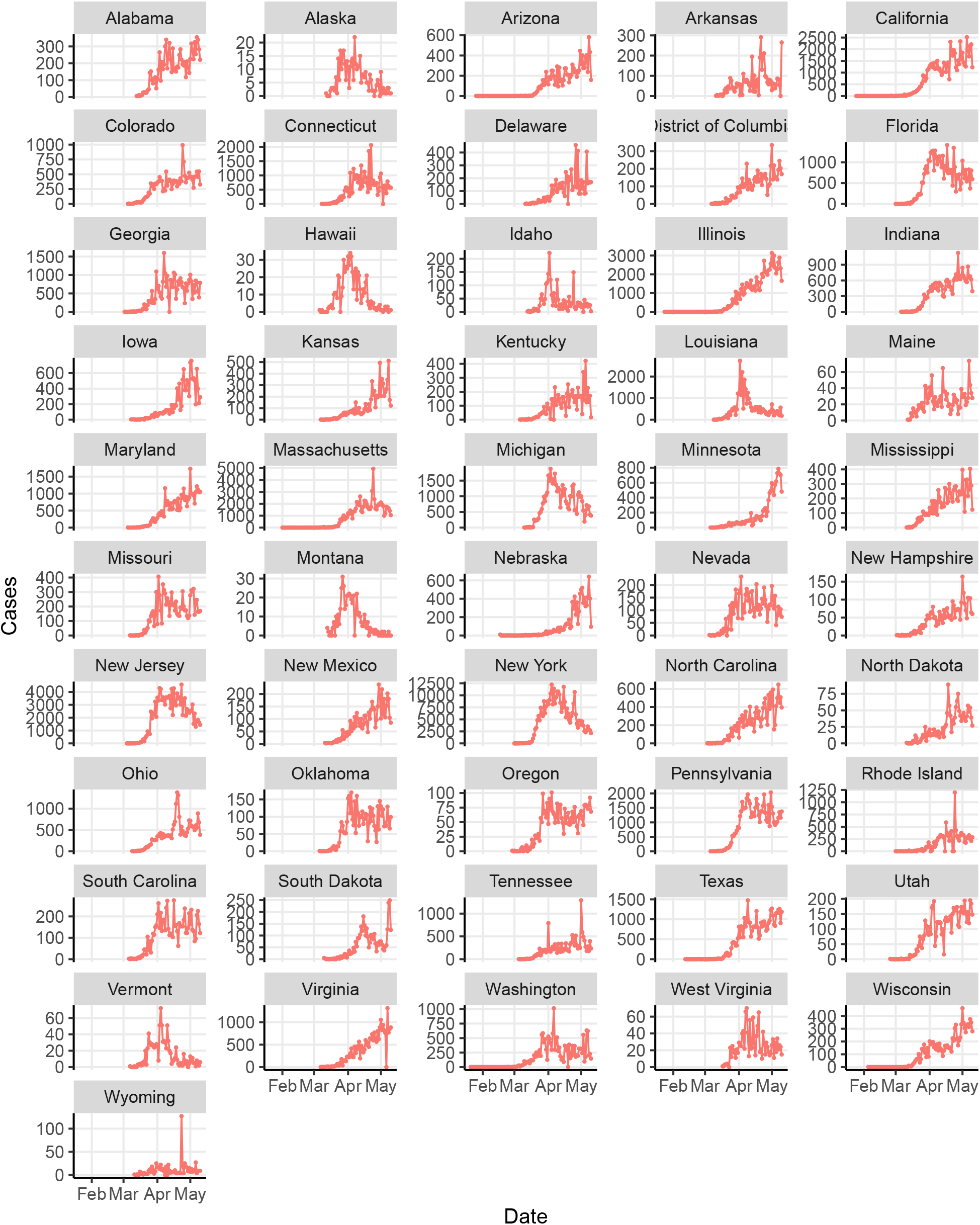
Daily counts of new cases by state in the U.S.

**Figure 8:**
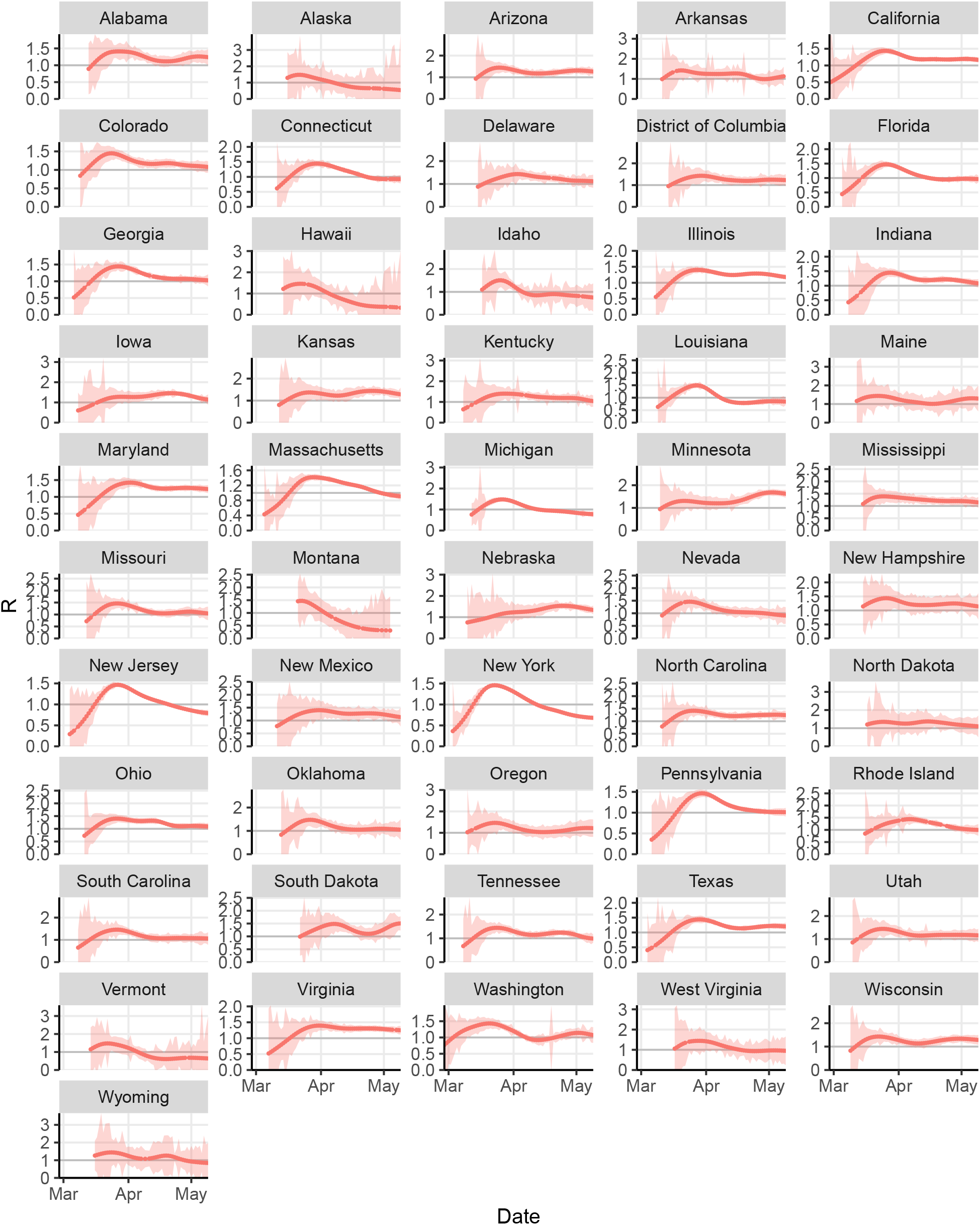
Estimated effective *R_t_* by state in the U.S.

**Figure 9:**
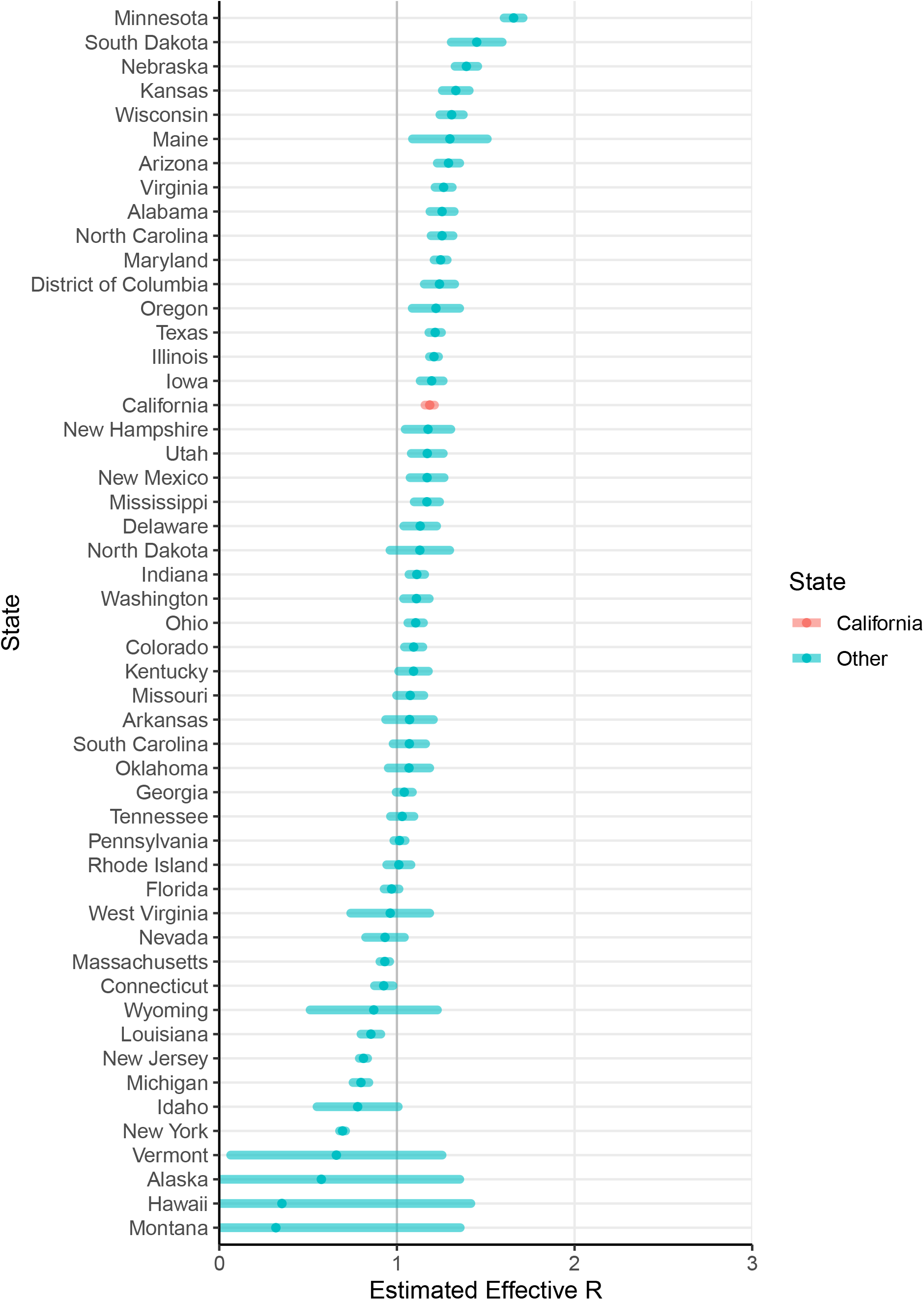
Most recent estimates of effective *R_t_* by state in the U.S., in descending order.

## Discussion

Our estimates confirmed that reproduction numbers appear to have declined in many locations where control measures such as sheltering at home have been adopted. Reproduction numbers appear to be widely variable across regions. As of May 9, 2020, our estimated effective reproduction number was highest in Minnesota, Nebraska, South Dakota, Kansas, and Arizona. Even in California, however, our estimates were indicative of supercriticality (*R_t_* > 1). Only New York, Michigan, New Jersey, and Louisiana were estimated to have achieved subcriticality by the data available on May 9, 2020.

The inclusion of substantial variability in the interval from symptom onset to case reporting in the serial interval distribution appears to cause the time series of estimated reproduction numbers to be highly smoothed, having lower peaks and higher troughs than when a less variable interval is assumed, and in comparison to other estimates (Lewnard et al. 2020; Darwin, Bandoy, and Weimer 2020; Donnat and Holmes 2020).

Because variablity in case reporting dates includes a substantial probability of negative serial intervals, in which a primary case may be reported after some of the cases it caused, the smoothing can include an increase in the reproduction number estimates occurring at later dates, as later cases are assigned some proportion of responsibility for earlier cases. The estimates at later dates may also be higher than expected as a result of the correction technique introduced by Cauchemez et al. (Cauchemez et al. 2006), which accounts for the portion of a case’s secondary cases not yet reported, when the serial interval extends beyond the last date reported. This effect is relatively large here, because our serial interval estimate includes long intervals.

Our analysis includes several limitations. Use of the Wallinga-Teunis estimator conventionally assumes complete reporting. Changes in reporting over time (such as inclusion of probable cases, or those resulting from increased or decreased testing) will yield biased estimates of *R_t_*, as would changes in reporting delays over time. Some jurisdictions have begun to report probable cases together with confirmed cases; such a change in the middle of a case series would yield an upward bias in the estimated *R_t_*.

Our estimates suggest that while control measures such as sheltering in place and social distancing appear to be helpful in reducing transmission, COVID-19 transmission continues to be a serious concern, as few states in the U.S. appear to have actually achieved subcriticality.

## Data Availability

All data used in this manuscript is publicly available from the New York Times and other sources, all linked in the manuscript.

## Appendix: Definition of Regions

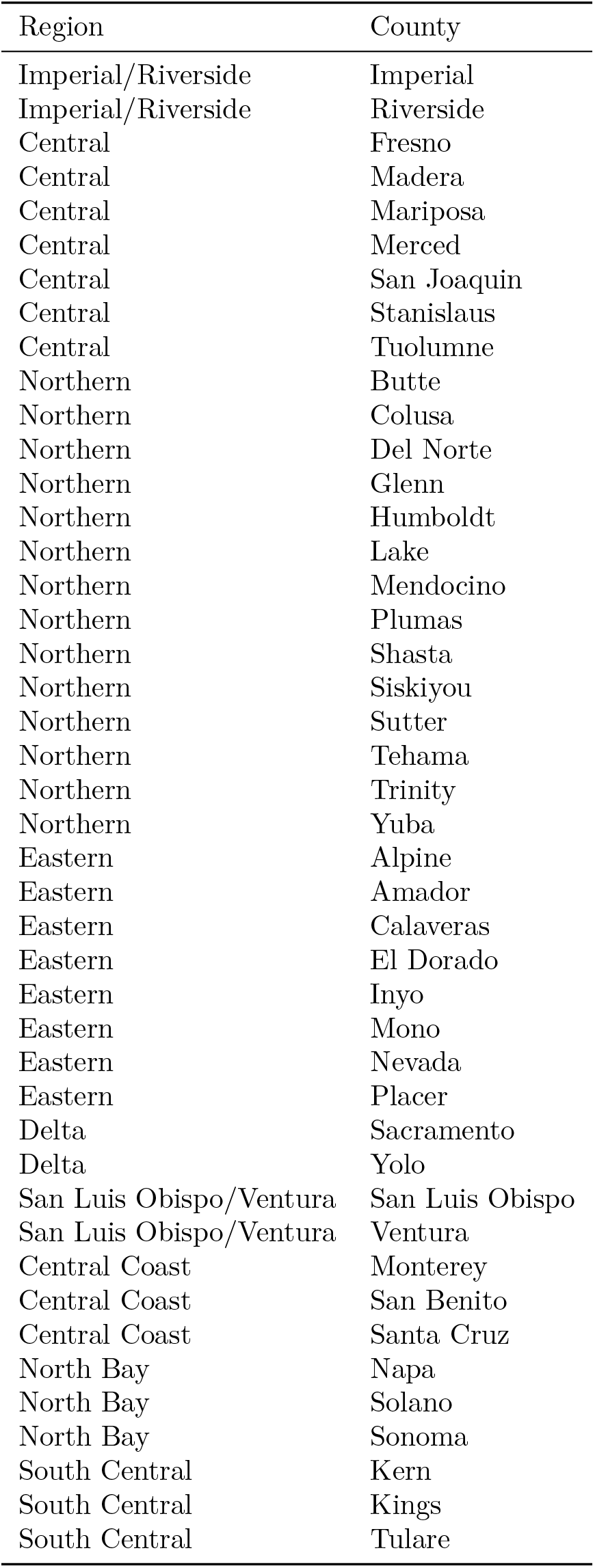

| Region | County |
| --- | --- |
| Imperial/Riverside | Imperial |
| Imperial/Riverside | Riverside |
| Central | Fresno |
| Central | Madera |
| Central | Mariposa |
| Central | Merced |
| Central | San Joaquin |
| Central | Stanislaus |
| Central | Tuolumne |
| Northern | Butte |
| Northern | Colusa |
| Northern | Del Norte |
| Northern | Glenn |
| Northern | Humboldt |
| Northern | Lake |
| Northern | Mendocino |
| Northern | Plumas |
| Northern | Shasta |
| Northern | Siskiyou |
| Northern | Sutter |
| Northern | Tehama |
| Northern | Trinity |
| Northern | Yuba |
| Eastern | Alpine |
| Eastern | Amador |
| Eastern | Calaveras |
| Eastern | El Dorado |
| Eastern | Inyo |
| Eastern | Mono |
| Eastern | Nevada |
| Eastern | Placer |
| Delta | Sacramento |
| Delta | Yolo |
| San Luis Obispo/Ventura | San Luis Obispo |
| San Luis Obispo/Ventura | Ventura |
| Central Coast | Monterey |
| Central Coast | San Benito |
| Central Coast | Santa Cruz |
| North Bay | Napa |
| North Bay | Solano |
| North Bay | Sonoma |
| South Central | Kern |
| South Central | Kings |
| South Central | Tulare |

## Footnotes

1 https://www.cdc.gov/media/releases/2020/s0126-coronavirus-new-cases.html, retrieved May 4, 2020.

2 https://www.cdph.ca.gov/Programs/OPA/Pages/NR20-006.aspx, retrieved May 4, 2020.

3 https://www.cdph.ca.gov/Programs/CID/DCDC/Pages/Immunization/nCoV2019.aspx, retrieved May 12, 2020.

4 https://covid-019.com/countries, retrieved May 12, 2020.

5 https://www.sfdph.org/dph/alerts/files/HealthOrderC19-07-%20Shelter-in-Place.pdf, http://www.acphd.org/media/561969/faqsorder-shelter-in-place-20200324.pdf, retrieved May 3, 2020.

6 https://covid19.ca.gov/img/Executive-Order-N-33-20.pdf, retrieved May 3, 2020.

7 https://sfmayor.org/article/san-francisco-issues-new-policy-face-coverings, retrieved May 3, 2020.

8 https://www.nytimes.com/interactive/2020/us/coronavirus-us-cases.html, retrieved May 4, 2020.

9 https://www.independent.com/2020/05/07/lompoc-prison-covid-19-cases-skyrocket-to-599/, retrieved May 10, 2020.

